# Simplified Perioperative Serplulimab and Chemotherapy for Resectable Squamous NSCLC: a Phase II Trial with Biomarker Analysis

**DOI:** 10.64898/2026.01.20.26344483

**Authors:** Fangqiu Fu, Haoxuan Wu, Chaoqiang Deng, Haiqing Chen, Qiangyuan Huang, Chongze Yuan, Xusheng Ding, Ting Ye, Yaodong Zhou, Sufeng Chen, Yihua Sun, Yawei Zhang, Jiaqing Xiang, Shengping Wang, Yuan Li, Bing Li, Yi Lu, Yang Zhang, Hong Hu, Haiquan Chen

## Abstract

**Purpose:** Squamous non-small cell lung cancer (sq-NSCLC) is a distinct subtype of NSCLC. This exploratory, phase II study investigated the feasibility and efficacy of a four-cycle perioperative regimen combining serplulimab with a taxane (paclitaxel or nab-paclitaxel) and carboplatin in patients with resectable stage II-IIIA sq-NSCLC.

**Methods:** This investigator-initiated, single-arm, phase II exploratory trial (NCT05775796) enrolled patients with histologically confirmed, resectable clinical stage II-IIIA squamous NSCLC. Patients received 2-3 cycles of neoadjuvant serplulimab plus taxane-carboplatin, followed by curative-intent surgery and 1-2 cycles of adjuvant treatment. The primary endpoint was major pathological response (MPR). Secondary endpoints included pathological complete response (pCR), R0 resection rate, overall response rate (ORR), safety, event-free survival (EFS), and overall survival (OS).

**Results:** A total of 30 patients without actionable driver mutations were enrolled and 29 underwent surgery. The median age was 65 years, and most were male smokers (n=28, 93.33%). An R0 resection was achieved in 28 patients (93.33%), and the MPR and pCR rates were 76.67% and 50.00%, respectively. Based on radiological assessments during the neoadjuvant phase, the ORR was 73.33% (95% CI: 54.11-87.72). Grade 3 and more treatment-related adverse events were predominantly hematologic and were generally manageable. Long-term EFS and OS data are not yet mature. Additionally, exploratory minimal residual disease analysis using circulating tumor DNA (ctDNA) in 27 patients showed a strong correlation between ctDNA clearance and pCR (*p*=0.004), suggesting ctDNA as a promising biomarker for immunochemotherapy response.

**Conclusions:** A four-cycle perioperative regimen of serplulimab combined with taxane-carboplatin demonstrated promising MPR and pCR rates with an acceptable safety profile in resectable sq-NSCLC patients. Long-term follow-up and future phase III trials are warranted to confirm survival benefits.

## 1. Introduction

Lung cancer is a leading cause of cancer incidence and mortality in the United States^1^. Over the past two decades, remarkable advances have been made in the diagnosis and treatment of non-small cell lung cancer (NSCLC), which accounts for over 80% of all lung cancers. The identification of actionable driver gene alterations has accelerated the development of targeted therapies, while the discovery of immune checkpoints has revolutionized NSCLC treatment. Squamous NSCLC (sq-NSCLC), constituting approximately 25-30% of NSCLC cases, is a distinct histological subtype characterized by low frequencies of actionable driver genomic alterations such as epidermal growth factor receptor (*EGFR*), anaplastic lymphoma kinase (*ALK*), and c-ros oncogene 1 (*ROS1*), leading to limited targeted therapeutic options^2, 3^. Immunotherapy targeting programmed cell death protein-1 (PD-1), or programmed death-ligand-1 (PD-L1) represents a promising anti-tumor strategy for sq-NSCLC. The randomized phase III KEYNOTE-407 trial demonstrated that pembrolizumab-based combination therapy significantly improved outcomes in metastatic sq-NSCLC patients across PD-L1 expression levels, with greater benefits observed in patients with higher PD-L1 expression^4^.

Early-stage (stage I-IIIA) NSCLC, representing approximately 40% of cases, is primarily treated with radical surgery^5^. However, recurrence or metastasis after surgery remains common. While perioperative chemotherapy improves prognosis, the absolute increase in 5-year survival rate is only about 5%^6, 7^. Recent landmark studies have highlighted the potential benefits of perioperative immunochemotherapy. In the CheckMate-816 trial, neoadjuvant nivolumab (an anti-PD-1 monoclonal antibody [mAb]) combined with chemotherapy resulted in a major pathological response (MPR) of 36.9% and pathological complete response (pCR) of 24.0% in resectable stage IB-IIIA NSCLC patients, with comparable safety to chemotherapy alone^8^. The subsequent KEYNOTE-671 trial further confirmed improved survival following perioperative pembrolizumab-based treatment^9^. Notably, subgroup analyses suggested enhanced benefit among patients with squamous histology, although dedicated studies specifically targeting sq-NSCLC in the perioperative setting remain limited.

Serplulimab, a fully humanized anti-PD-1 mAb, has demonstrated higher affinity for the PD-1 receptor and greater anti-tumor activity compared to nivolumab and pembrolizumab^10^. Its efficacy in lung cancer has been validated by the ASTRUM-005 and ASTRUM-004 trials, showing improved outcomes with serplulimab-based therapy in extensive-stage small-cell lung cancer (ES-SCLC) and advanced PD-L1-positive sq-NSCLC, respectively^3, 11^. Building upon these findings, we have initiated a single-center, single-arm, exploratory phase II clinical trial to evaluate the feasibility and preliminary efficacy of a simplified, four-cycle perioperative regimen combining serplulimab with chemotherapy in patients with resectable stage II-IIIA sq-NSCLC. Additionally, we explored circulating tumor DNA (ctDNA)-based minimal residual disease (MRD) as a potential predictive biomarker for treatment response.

## 2 Methods

### 2.1 Study Design and Participants

This investigator-initiated, single-center, single-arm phase II exploratory clinical trial (NCT05775796) planned to enroll 30 patients with histologically or cytologically confirmed, resectable clinical stage II-IIIA sq-NSCLC. It aimed to evaluate the feasibility of a perioperative regimen consisting of four cycles of serplulimab combined with taxane and platinum-based chemotherapy (Supplementary Figure 1). Key eligibility criteria are summarized in Supplementary Table 1, with full eligibility details provided in the study protocol.

Eligible participants received a total of four cycles of perioperative serplulimab-based treatment, comprising two to three neoadjuvant cycles and one to two postoperative adjuvant cycles. After treatment completion, patients were followed up every three months until death. The primary endpoint was MPR. Secondary endpoints included pCR, R0 resection rate, overall response rate (ORR), event-free survival (EFS), overall survival (OS), and safety.

This study was conducted in accordance with the Declaration of Helsinki and the International Conference on Harmonisation Good Clinical Practice Guidelines (ICH-GCP). The study protocol and amendments were reviewed and approved by the Clinical Research Ethics Committee of Fudan University Shanghai Cancer Center (Approval No: 2211264-18), and written informed consent was obtained from all participants.

### 2.2 Treatment and Procedures

All enrolled patients in the study received perioperative serplulimab (fixed dose, 300 mg intravenously [i.v.] every three weeks [Q3W]), combined with paclitaxel (175 mg/m²) or nab-paclitaxel (260 mg/m²) and carboplatin (area under the curve [AUC] =5). Treatment consisted of two to three cycles administered preoperatively (neoadjuvant phase), followed by curative-intent surgery and one to two postoperative cycles (adjuvant phase). The exact number of cycles was adjusted based on imaging-based tumor response, clinical tolerability, and investigator judgment. Detailed treatment regimens and procedures are described in the study protocol. Patients were followed up every three months post-treatment.

### 2.3 Study Endpoints and Assessment

The primary endpoint was the postoperative MPR rate, defined as the proportion of patients with ≤10% viable tumor cells remaining in the primary tumor bed after neoadjuvant therapy, regardless of residual tumor cells in lymph nodes. Key secondary endpoints included the R0 resection rate and the pCR rate. An R0 resection required fulfillment of specific criteria established by the International Association for the Study of Lung Cancer (IASLC) Staging and Prognostic Factors Committee, including microscopically negative resection margins, a systematic lymph node dissection covering at least six lymph node stations (minimum three intrapulmonary/hilar and three mediastinal stations), no extracapsular nodal involvement, and no involvement of the highest resected lymph node^12^. The pCR rate was defined as the proportion of patients with 0% residual viable tumor cells in both the primary tumor bed and resected lymph nodes after neoadjuvant therapy. Pathological response was assessed by two experienced senior pulmonary pathologists from the Department of Pathology at Fudan University Shanghai Cancer Center who were blinded to the treatment assignment.

Secondary endpoints also included tumor response during the neoadjuvant phase, EFS, OS, and the safety profile. Tumor response was assessed by investigators according to RECIST v1.1, with ORR defined as the proportion of patients achieving a partial response (PR) or complete response (CR) after neoadjuvant therapy. EFS was defined as the time from enrollment to disease progression precluding surgery, local or distant recurrence, or death from any cause. OS was defined as the time from enrollment to death from any cause. Safety assessments were conducted according to the National Cancer Institute Common Terminology Criteria for Adverse Events (CTCAE) version 5.0. Adverse events (AEs) were recorded throughout the treatment and follow-up periods. This study also planned to explore the efficacy differences between clinical stage II and III patients to further evaluate the feasibility of the perioperative regimen.

### 2.4 DNA Sequencing and MRD Detection

Additionally, pre-treatment tumor tissue samples were collected for whole exome sequencing (WES) to identify gene mutations in sq-NSCLC. Specifically, only samples containing ≥30% tumor cells were included in the analysis. The MagPure FFPE DNA/RNA LQ kit (Magen, Guangzhou, China) was used to extract genomic DNA (gDNA) from formalin-fixed, paraffin-embedded (FFPE) tumor tissues. Peripheral blood gDNA was extracted with the MagPure Universal DNA Kit (Magen, Guangzhou, China). DNA samples meeting quality criteria were subjected to WES library preparation, exome capture, and quantification. Sequencing was performed on a NovaSeq 6000 platform (Illumina, San Diego, CA, USA) with paired-end reads, targeting an average coverage of approximately 500× for tumor samples and 150× for matched normal samples^13^.

Peripheral blood samples were collected at predefined time points using Cell-Free DNA BCT tubes (Streck, La Vista, NE, USA). Cell-free DNA (cfDNA) was isolated using the QIAamp Circulating Nucleic Acid Kit (Qiagen) and analyzed using the PROPHET assay to detect MRD^13^. High-depth sequencing, utilizing unique molecular identifier (UMI)-based technology, was conducted on the Illumina NovaSeq 6000 platform (San Diego, CA, USA) with paired-end reads, targeting a raw sequencing depth of 100,000× for personalized panels.

After adapter trimming and quality control, single nucleotide variants (SNVs) and insertions/deletions (INDELs) with at least 5 reads and a variant allele frequency (VAF) of ≥3% were identified. Germline mutations were filtered by comparing the fold change in VAF between tumor and matched normal samples, excluding variants with a fold change <3 or both VAFs >10%. Variant annotation was conducted using ANNOVAR and SnpEff v3.6. Up to 50 high-priority variants with a VAF ≥3.0% were selected, excluding those in repetitive, high GC (>75%), or homologous regions. A personalized biotinylated capture probe pool was created, and circulating tumor DNA (ctDNA) was sequenced using this custom panel. Somatic mutation significance was calculated using a Poisson distribution by the PROPHET assay^13^, which tracks up to 50 patient-specific variants. Utilizing hybrid capture with UMIs, the assay achieves an average raw sequencing depth of approximately 100,000×. The analytical limit of detection (LOD) was established via serial dilutions of cancer cell lines and reference standards, yielding 0.0036% at 20ng DNA input and 0.0018% at 60ng. Consequently, the validated assay LOD was set at 0.004% with a specificity exceeding 99%. Accordingly, ctDNA clearance was defined as levels below this validated LOD. Mutations with *p* <0.05 were considered significant, and MRD-positive status was defined by the presence of at least two significant mutations with *p* <0.005.

### 2.5 Statistical Analysis

This single-arm study employed a statistical hypothesis based on historical data. According to previous research, the MPR rate for neoadjuvant chemotherapy was approximately 9%^8^. Assuming an anticipated MPR rate of 27%, with a two-sided alpha of 0.05, a power of 80%, and a dropout rate of 10%, the calculated sample size was 30 patients. Three analysis sets were defined for statistical analysis: the full analysis set (FAS), including all patients who received at least one dose of study treatment and had at least one post-treatment assessment; the per-protocol set (PPS), a subset of FAS excluding patients with major protocol deviations; and the safety set (SS), including all patients who received at least one dose of study treatment with available safety data.

Statistical analyses were performed using R software (version 4.3.2). Analyses comprised: (1) a summary of patient enrollment, exclusions, withdrawals, and completion status; (2) descriptive summaries of baseline demographic, clinical, and treatment characteristics; (3) assessment of treatment efficacy for primary (MPR) and secondary endpoints (pCR, R0 rate, ORR, EFS, OS); and (4) safety evaluations, including treatment-related adverse events (TRAEs). Continuous variables were described using means, standard deviations (S.D.), medians, and ranges. Categorical variables were summarized using counts and percentages. The 95% confidence intervals (CIs) for ORR were calculated using the Clopper-Pearson method. EFS and OS were estimated using the Kaplan-Meier method. To compare differences between subgroups, the independent samples t-test or paired samples t-test was used for normally distributed continuous variables, the Mann-Whitney U test or Kruskal-Wallis H test was used for non-normally distributed continuous variables, and the Fisher’s exact test was used for categorical variables. Additionally, logistic regression analysis was conducted in this study based on the outcome variable to evaluate the associations between potential influencing factors and the outcome. All statistical tests were two-sided, and a *p*-value of less than 0.05 was considered statistically significant.

## 3. Results

### 3.1 Study Overview, Baseline Characteristics, and Treatment

Between April 2023 and February 2024, 30 patients with stage II–IIIA sq-NSCLC were enrolled and received at least one dose of the study treatment. All 30 patients were Asians and included in the FAS (Figure 1). Due to safety events, two patients received only one cycle of neoadjuvant therapy, with one patient subsequently not undergoing definitive surgery. Among the remaining 28 patients who completed the protocol-specified neoadjuvant treatment, 19 received two cycles and 9 received three cycles. Twenty-nine patients underwent definitive surgery, of whom six declined subsequent adjuvant consolidation therapy, while the remaining 23 completed the full perioperative regimen as planned.

**Figure 1.**
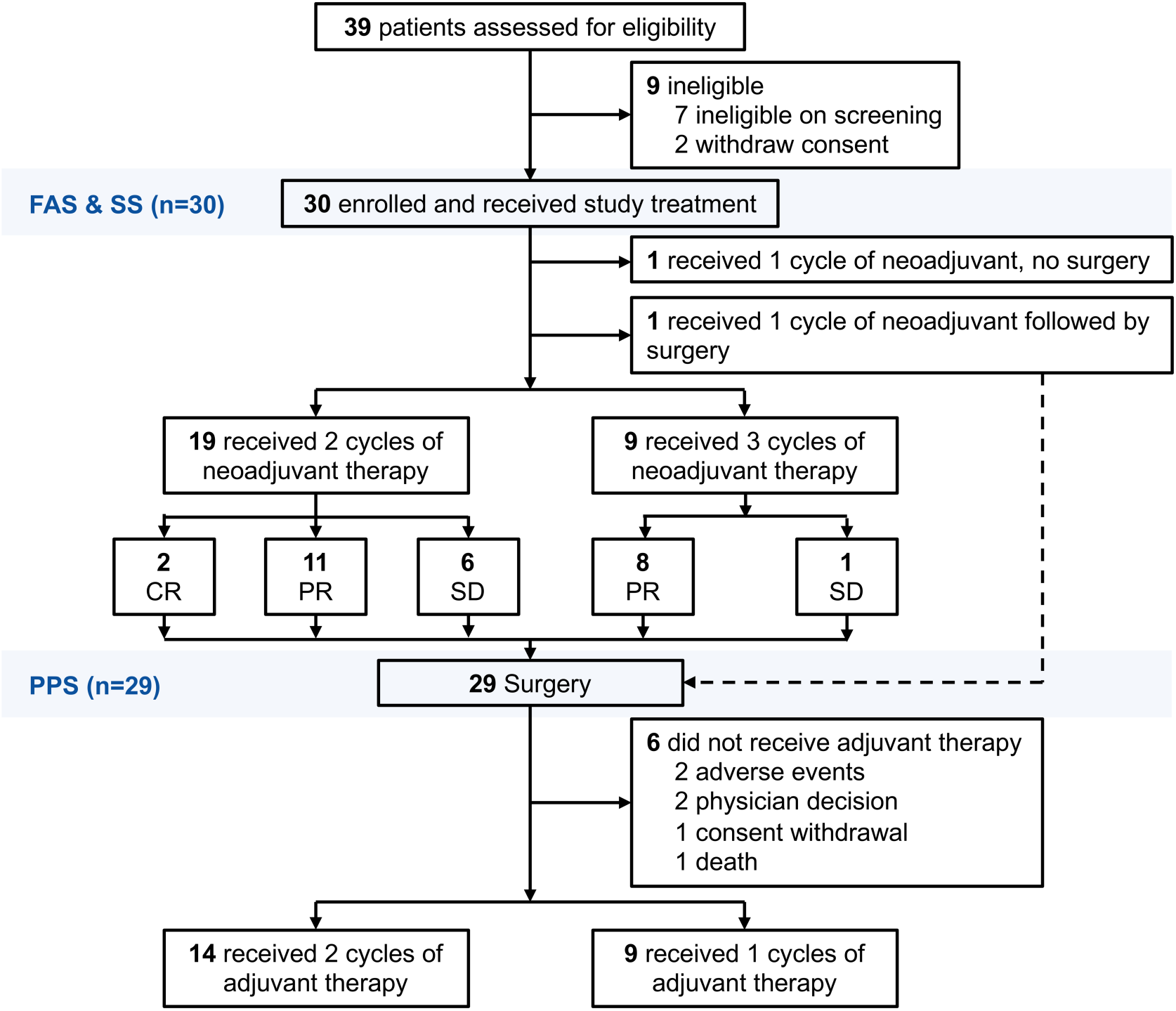
Trial profile. FAS, full analysis set; SS, safety set; PPS, per-protocol set; CR, complete response; PR, partial response; SD, stable disease.

Baseline and treatment characteristics are summarized in Table 1. The median age was 65 years (range: 35-75), and most patients were male smokers (n=28, 93.33%). Fourteen patients (46.67%) had an Eastern Cooperative Oncology Group performance status (ECOG PS) of 0. Nearly half of the patients presented with stage II disease (2 with stage IIA and 12 with stage IIB), while the remaining 16 patients had stage IIIA disease; 8 patients (26.67%) had N2 lymph node involvement. PD-L1 expression (tumor proportion scores [TPS] ≥1%) was detected in 24 patients (80.00%), and 12 patients (40.00%) demonstrated high PD-L1 expression (TPS ≥50%). Twenty-two patients (73.33%) received nab-paclitaxel plus carboplatin, and the remaining 8 patients (26.67%) received paclitaxel plus carboplatin. Among the 29 patients who completed surgery, the median interval from the last neoadjuvant dose to surgery was 35 days (Table 2). All patients (29/29, 100%) underwent muscle-sparing thoracotomy, with lobectomy being the most common surgical approach (n=17, 58.62%). All patients underwent complete mediastinal lymph node dissection, with a mean of 19.79 lymph nodes removed.

**Table 1.**
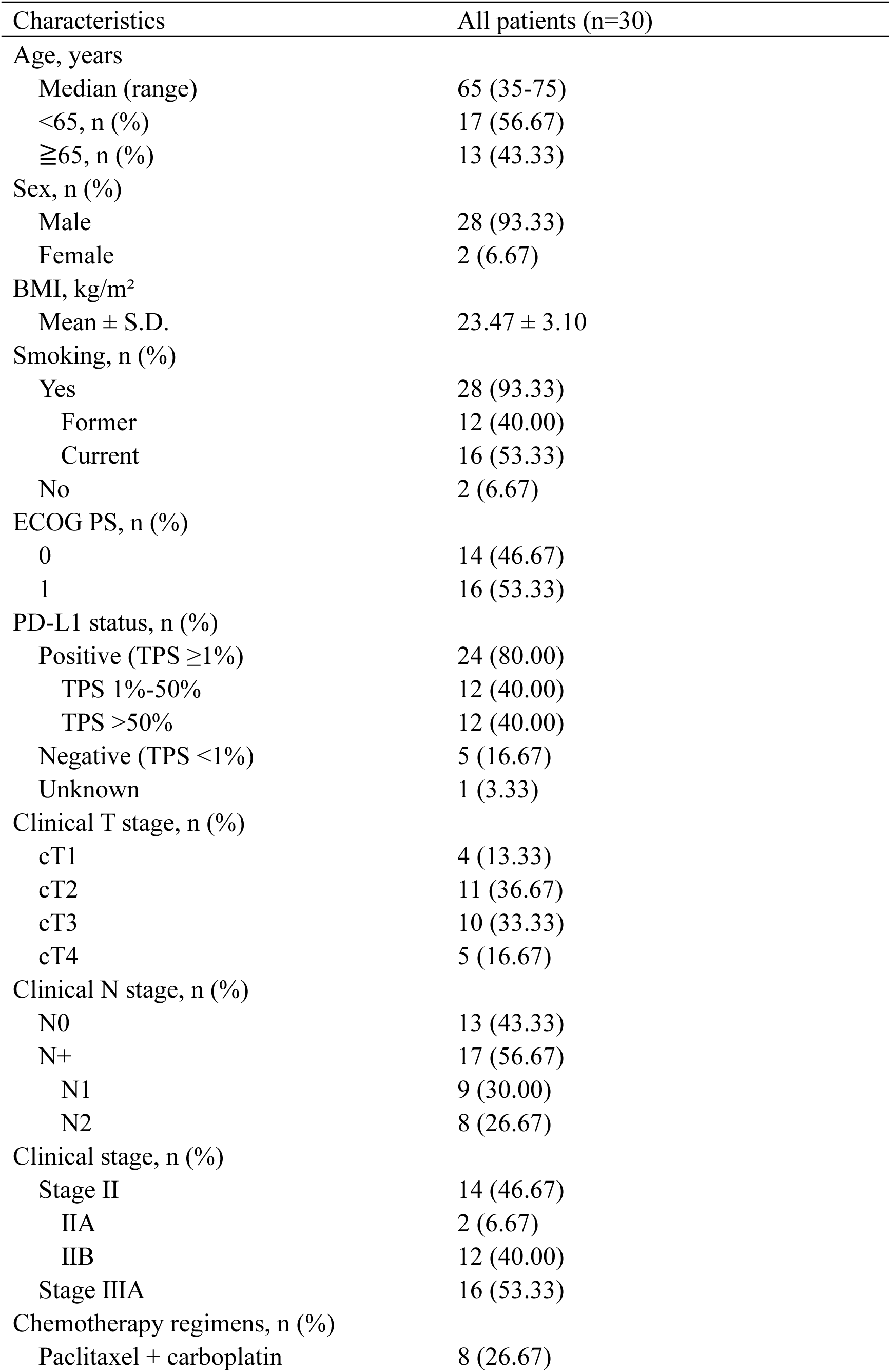
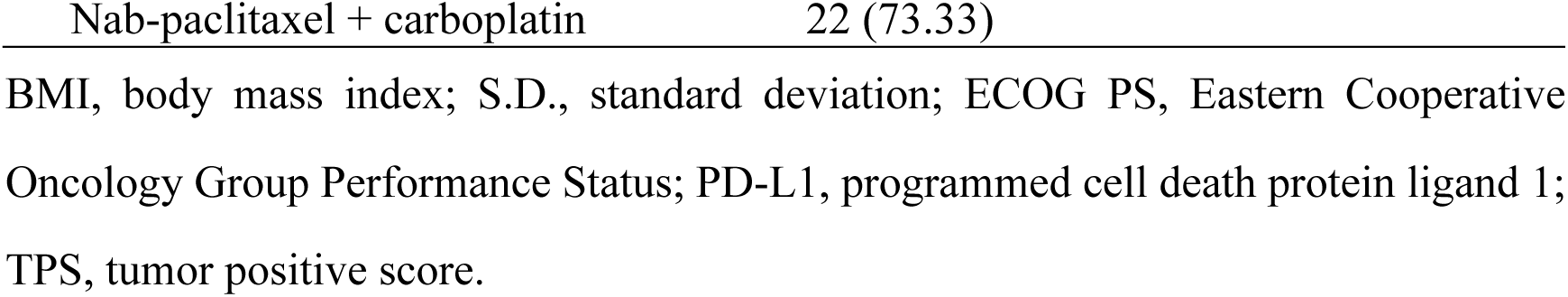
Baseline Characteristics.

**Table 2.**
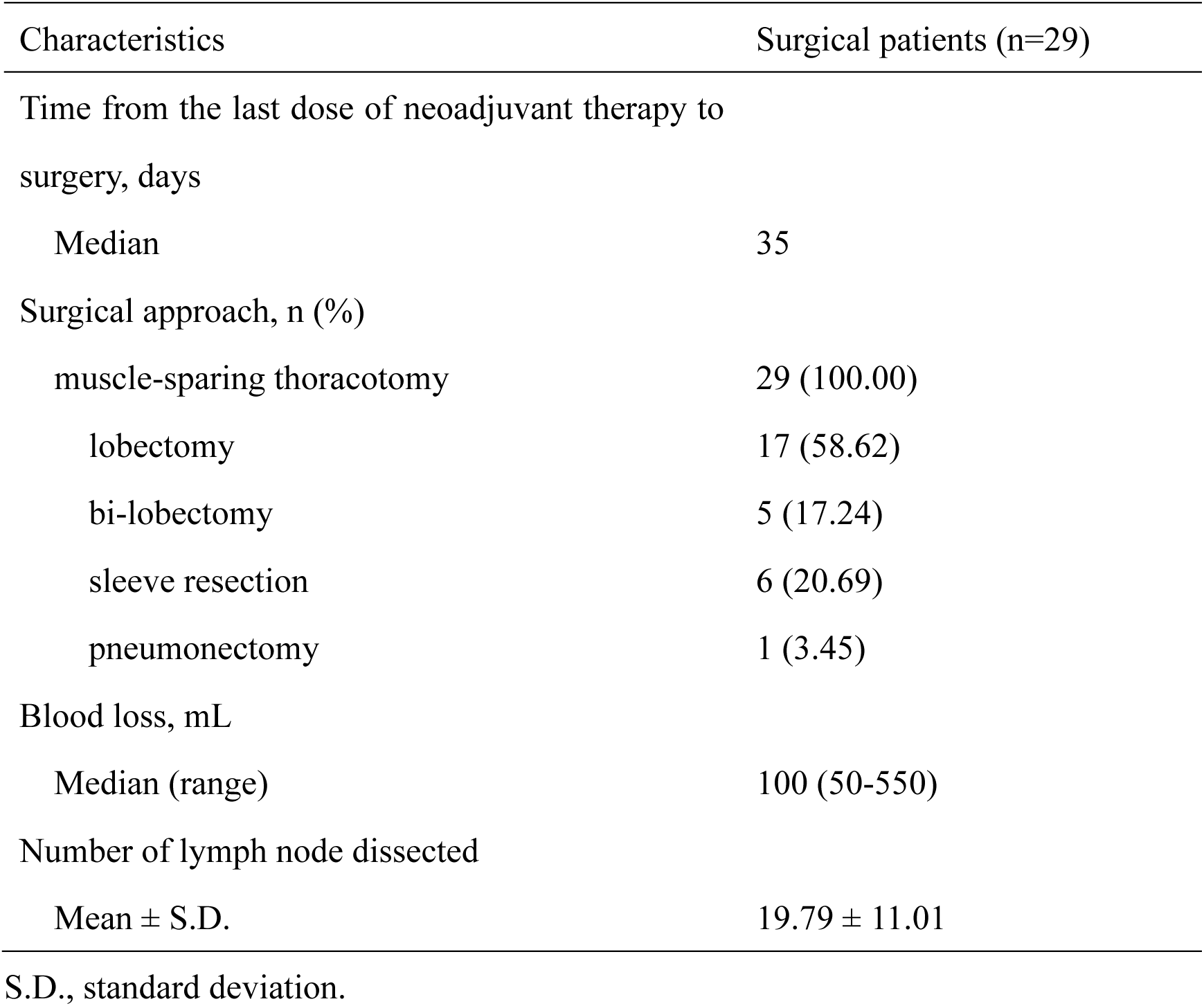
Surgery Characteristics.

By the data cutoff in November 2024, all patients had completed the study treatment, with a median follow-up of 13.6 months (IQR 11.49-15.86). Four patients experienced EFS events, including two with recurrence and two deaths. Survival data remain immature, with median EFS and OS not yet reached (Supplementary Figures 2 and 3).

### 3.2 Antitumor Activity of Serplulimab-Based Neoadjuvant Treatment

In the intention-to-treat (ITT) population (n=30), the ORR during neoadjuvant treatment was 73.33% (22/30, 95% CI: 54.11-87.72) with no progressive disease (PD) (Table 3). The 29 patients who underwent curative surgery were included in the PPS as the primary efficacy analysis population. Twenty-eight achieved R0 resection (R0 resection rate: 96.55% in the PPS; 93.33% in the FAS), while one patient had R2 resection. Postoperative pathological evaluation revealed that 23 patients achieved MPR, and 15 patients achieved pCR, resulting in MPR and pCR rates of 76.67% (95% CI: 57.72-90.07) and 50.00% (95% CI: 31.30-68.70) in the ITT population (79.31% and 51.72% in the PPS, respectively). Notably, 11 patients with PR and two with SD as their best radiological responses achieved pCR post-surgery (Figure 2). These discrepancies between radiographic and pathological assessments are potentially due to immune-related tumor dynamics requiring further investigation. Additionally, postoperative pathology showed negative lymph node involvement in 23 patients (79.31%), including five with baseline N2 disease. Subgroup analysis demonstrated no significant differences in MPR or pCR rates based on clinical stage (II vs. III) or neoadjuvant cycles (one vs. two vs. three) (Table 4), and the logistic regression analysis also showed no significant associations (Supplementary Table 2). The findings in the ITT population were consistent with those in the PPS (Supplementary Tables 3 and 4). A post-hoc analysis assessed changes in tumor biomarkers before and after neoadjuvant therapy, including serum concentrations of carcinoembryonic antigen (CEA), squamous cell carcinoma antigen (SCC Ag), neuron-specific enolase (NSE), cancer antigen 125 (CA125), and cytokeratin-19 fragments (CYFRA21-1). All biomarkers showed a significant decrease following neoadjuvant therapy (all *p*-value <0.05) (Supplementary Figure 4, Supplementary Table 5). Additionally, patients who achieved radiological responses (CR and PR) and significant reductions in serum biomarkers were more likely to achieve MPR (all *p*-value <0.05).

**Figure 2.**
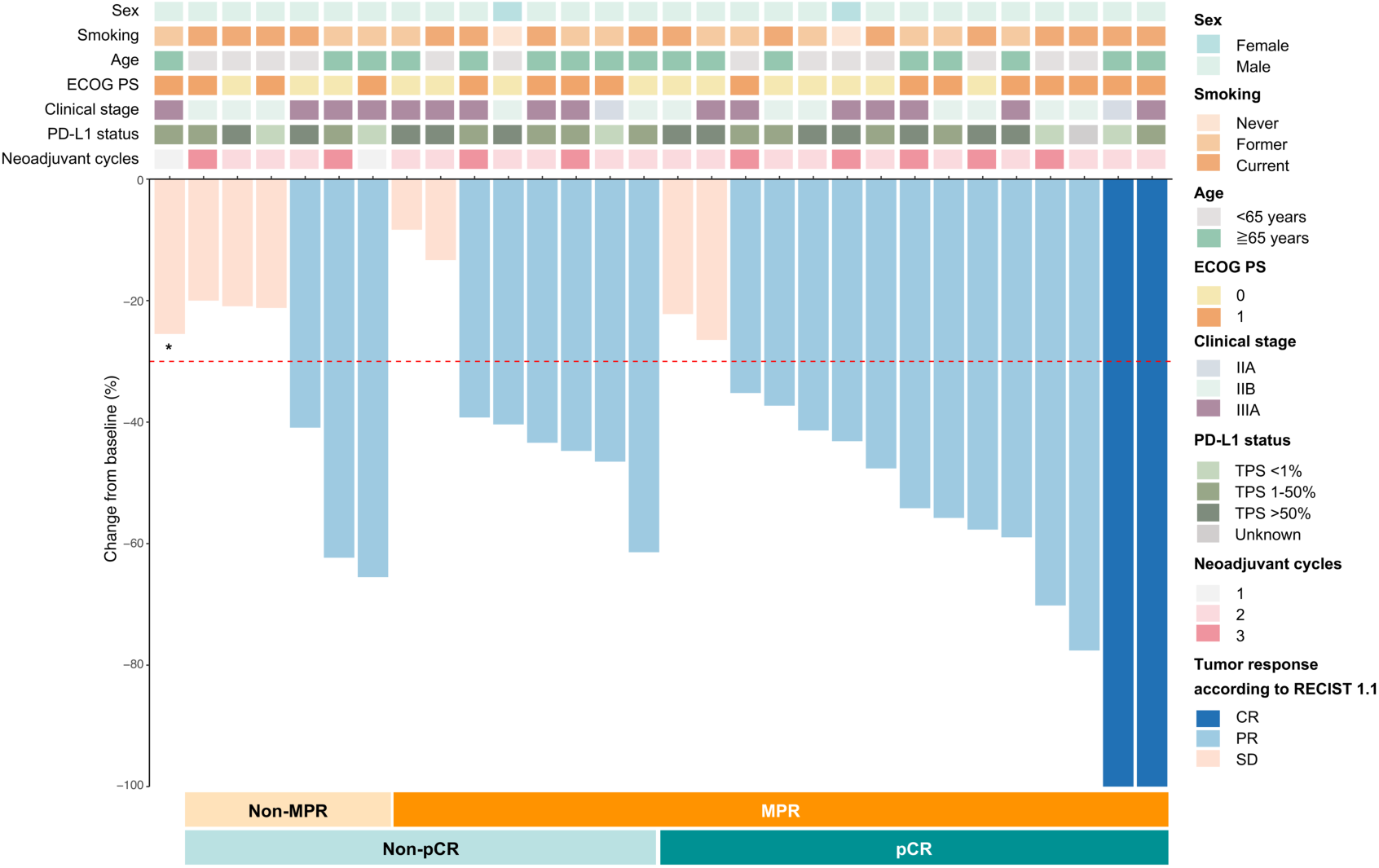
Tumor response. *Patient who achieved SD after one cycle of neoadjuvant therapy and did not undergo the planned surgery. ECOG PS, Eastern Cooperative Oncology Group Performance Status; PD-L1, programmed cell death protein ligand 1; TPS, tumor positive score; CR, complete response; PR, partial response; SD, stable disease; MPR, major pathological response; pCR, pathological complete response; RECIST 1.1, Response Evaluation Criteria in Solid Tumors version 1.1.

**Table 3.**
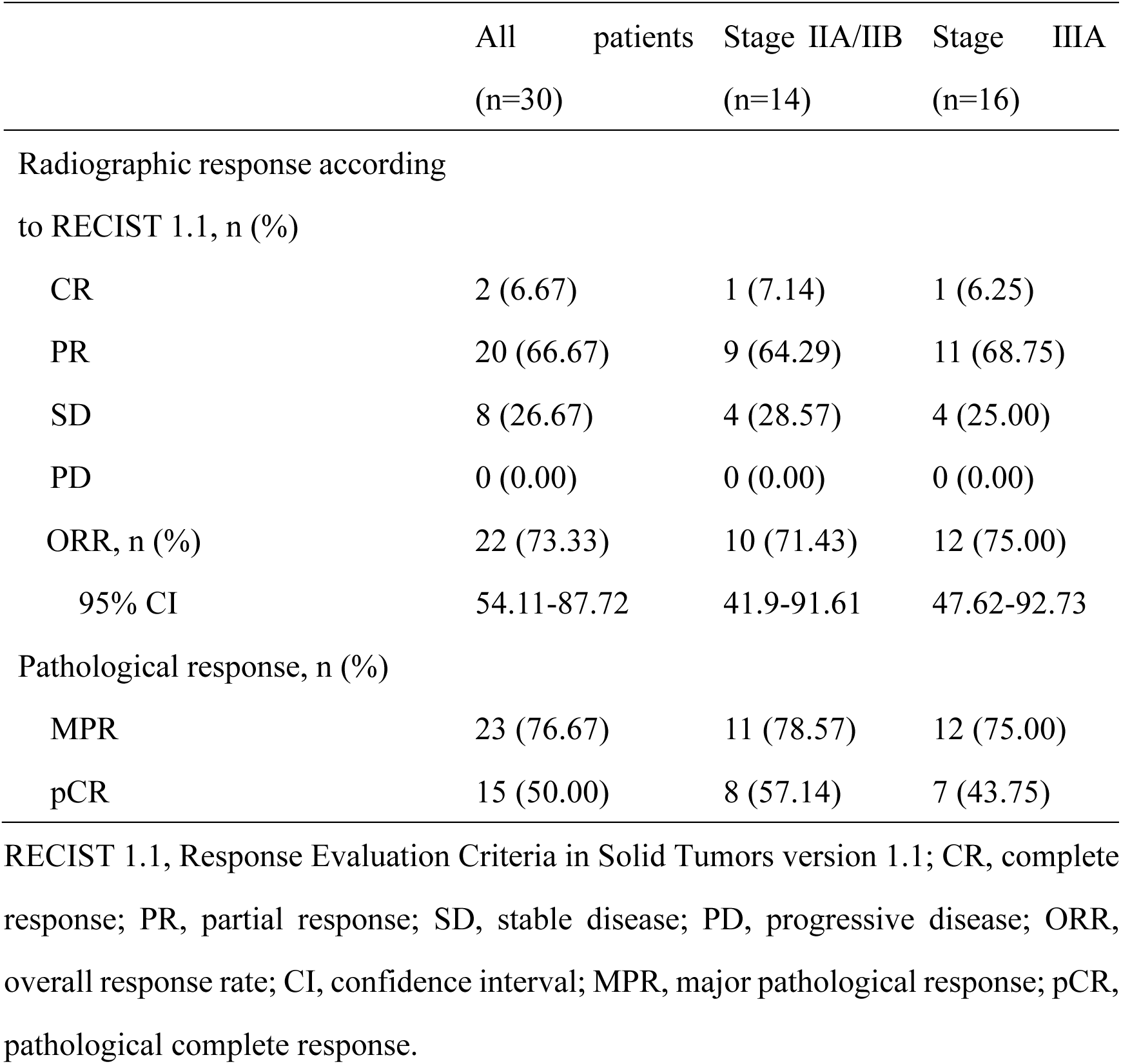
Tumor Response of Patients in the Intention-to-Treat Population.

**Table 4.**
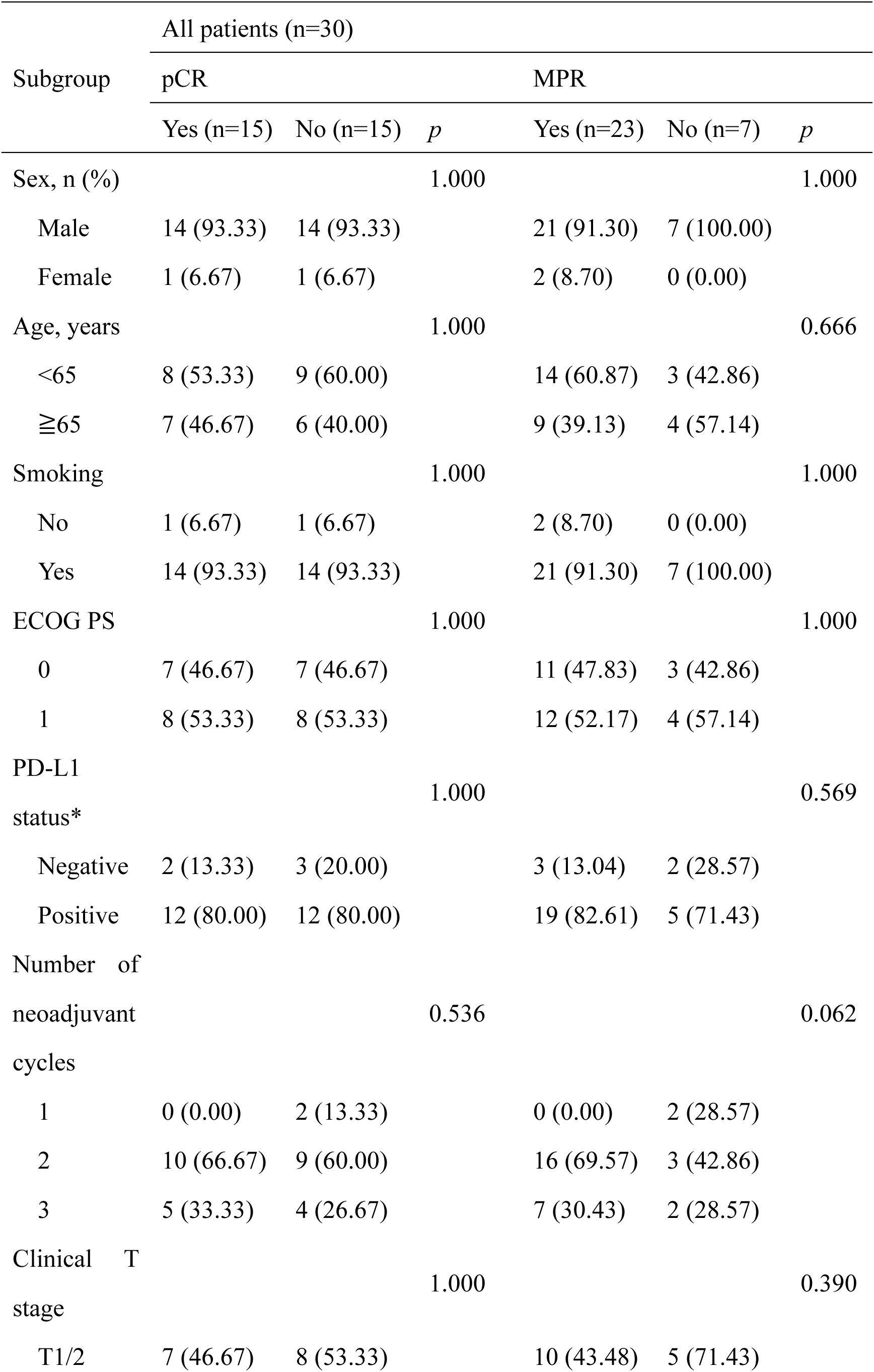
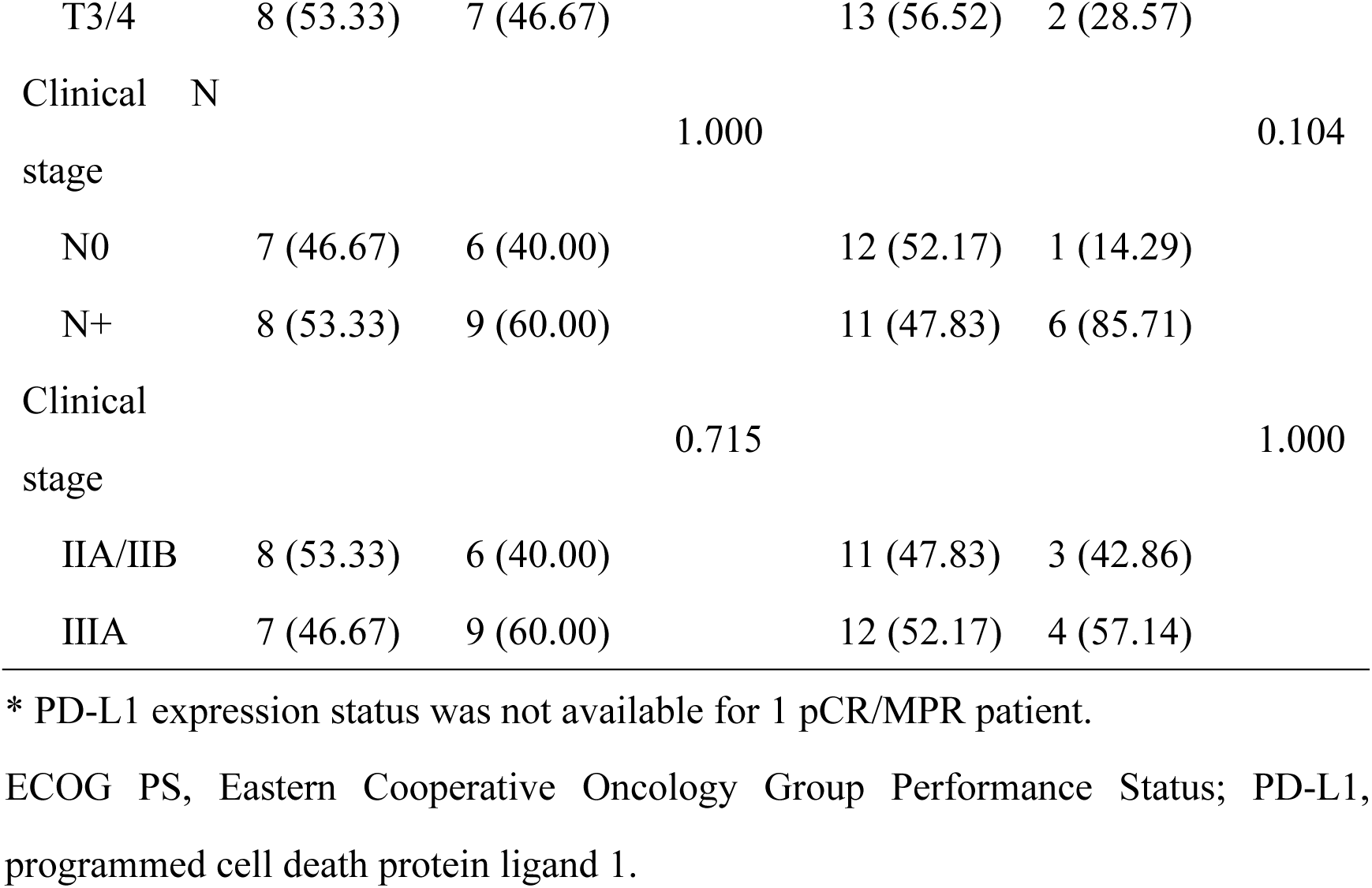
Subgroup Analysis of pCR and MPR in the Intention-to-Treat Population All patients (n=30)

### 3.3 Safety Profile

Generally, the TRAEs from this trial was manageable. All patients experienced at least one TRAE, and the incidence of grade ≥3 TRAEs was 26.66%. The most common TRAE was hematologic, including anaemia and neutropenia. Neutropenia was the most common grade ≥3 TRAEs, accounting for 23.33% of patients (Supplementary Table 6).

### 3.4 Predicting pCR After Immunochemotherapy Using MRD

It is widely recognized that pCR serves as a surrogate marker of improved clinical outcomes and is frequently used as a key endpoint in clinical trials^14^. Consequently, accurately predicting pCR status following immunochemotherapy is of great importance. In our study, tumor-informed MRD assays were successfully performed in 27 patients. Blood samples were collected at multiple time points: before the neoadjuvant phase (Time Point [TP]-0), prior to surgical resection (TP-1), at the start of the adjuvant phase (TP-2), and during follow-up (TP-3, TP-4, etc.) (Figure 3A). By the data cutoff, a total of 97 blood samples had been analyzed for MRD. Baseline WES revealed that *TP53* was the most common mutation, detected in 93% of patients (Figure 3B). At baseline (TP-0), all patients were ctDNA-positive (Figure 3C). Following immunochemotherapy, MRD negativity was achieved in 52% of patients, while 48% remained MRD-positive (Figure 3C). Most patients showed a decline in ctDNA ratios after immunochemotherapy, though a subset displayed increased ctDNA levels during follow-up (Figure 3D).

**Figure 3.**
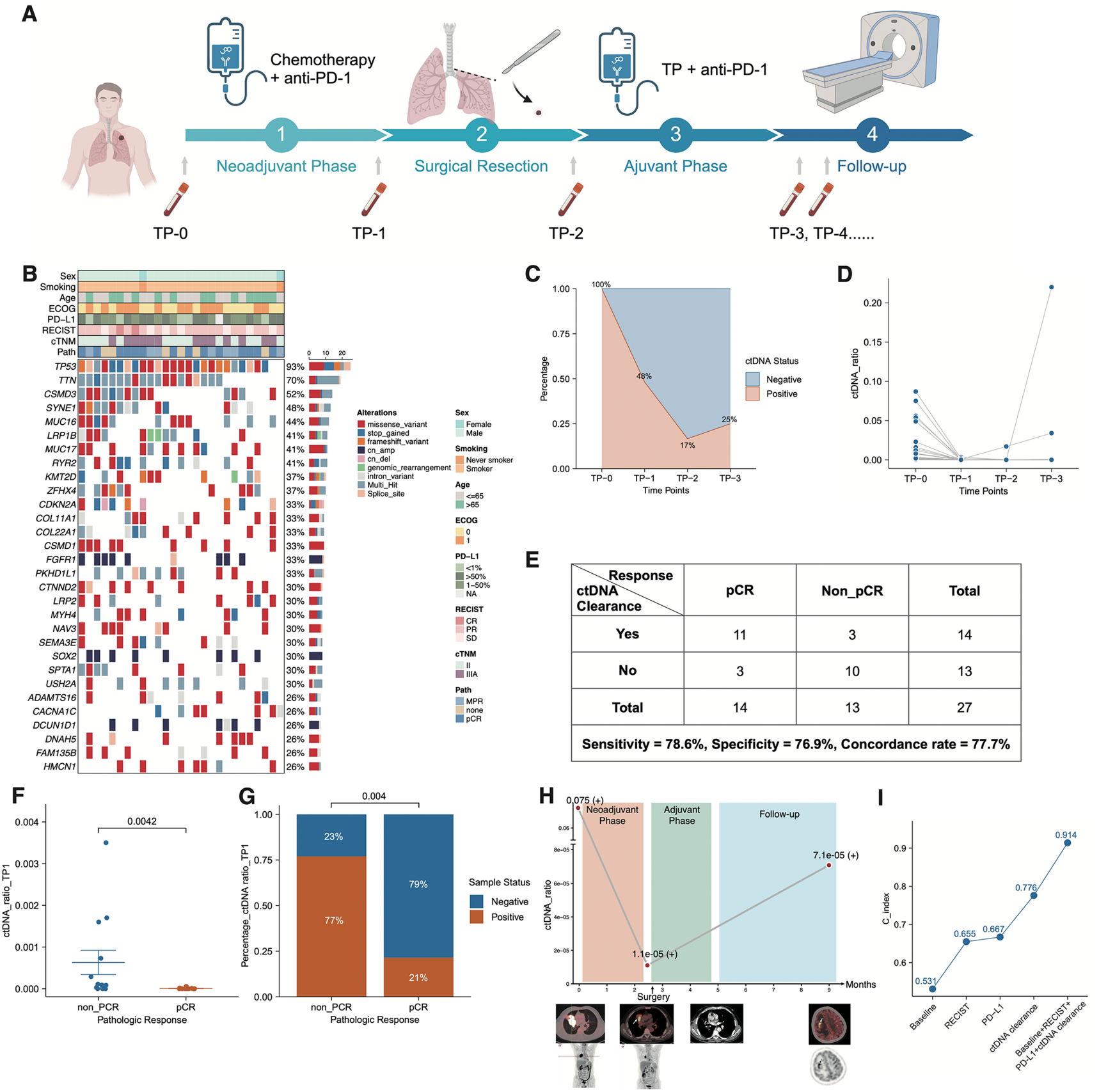
The association between ctDNA levels and pCR. (A) Schematic representation of the study design, including treatment timeline and blood sample collection points. (B) Heatmap showing the top 30 most frequently altered genes in the cohort. (C) Bar plots illustrating the percentage of patients with positive ctDNA at various time points. (D) Graphs depicting the dynamic changes in ctDNA ratios across different time points. (E) Performance of ctDNA clearance after treatment in predicting pathological response. (F) Comparison of ctDNA ratios between non-pCR and pCR patients. (G) Bar plots revealing the percentage of patients with positive ctDNA in patients with non-pCR and pCR. (H) Representative case (LC01002) demonstrating dynamic changes in ctDNA ratio alongside radiological findings. (I) Comparative C-index values of different predictive models for pCR in sq-NSCLC patients undergoing immunochemotherapy.

Further analyses showed that ctDNA clearance could accurately predict pCR status with a sensitivity of 78.6% and a specificity of 76.9% (Figure 3E). Patients who achieved pCR had significantly lower mean ctDNA ratios compared to those in the non-pCR group (*p*=0.0042) (Figure 3F). Of the non-pCR patients, 77% remained MRD-positive, whereas only 21% of pCR patients exhibited positive MRD status (*p*=0.004) (Figure 3G). Interestingly, one patient (LC01002) achieved pCR after neoadjuvant immunochemotherapy but retained a positive MRD status (ctDNA ratio: 1.1E-5) (Figure 3H). Six months post-surgery, this patient developed brain metastases, suggesting that ctDNA clearance may serve as a more sensitive indicator of residual tumor burden than pCR alone. We next compared the predictive capabilities of baseline characteristics, RECIST response, PD-L1 expression, ctDNA clearance, and a combined model for determining pCR status. While ctDNA clearance alone achieved a C-index of 0.776, outperforming both RECIST response (C-index: 0.655) and PD-L1 expression (C-index: 0.667), the integrated model incorporating all variables achieved a C-index of 0.914 (Figure 3I). Taking together, these findings underscore the potential of ctDNA clearance as a robust predictor of pCR in resectable sq-NSCLC, which may need future validation

## 4. Discussion

Several landmark trials have established the benefits of perioperative immunochemotherapy in resectable NSCLC. The CheckMate-816 trial demonstrated that neoadjuvant nivolumab combined with chemotherapy significantly improved the pCR rates ( 24.0% vs. 2.2%), prolonged EFS (median, 43.8 vs. 18.4 months; HR [95% CI]: 0.66 [0.49–0.90]), and increased the 4-year OS rates (71% vs. 58%)^15^, and the findings validated among Asian populations^16, 17^. Notably, CheckMate-816 allowed optional postoperative adjuvant therapy. Subsequently, the KEYNOTE-671 trial advanced the perioperative immunotherapy approach by combining neoadjuvant pembrolizumab and chemotherapy with mandatory adjuvant pembrolizumab, resulting in significantly increased pCR rate from 4.0% to 18.1% and extended the median EFS to 47.2 months (vs. 18.3 months for the control group; HR [95% CI]: 0.59 [0.48-0.72])^9^. Despite these breakthroughs, most studies focused primarily on comparing immunotherapy agents rather than refining treatment strategies or individualizing therapy^18–20^. Our study is the first to specifically examine the feasibility of a perioperative immunochemotherapy regimen in resectable stage II-IIIA squamous NSCLC, using serplulimab combined with a taxane plus carboplatin. Uniquely, we compressed the perioperative regimen to only four cycles in total, either two or three cycles of neoadjuvant therapy, followed by one or two postoperative cycles based on radiographic response and patient preference. This flexibility may significantly maximize patient compliance and reduce treatment burden. However, it might also introduce variability, limiting the interpretability and generalizability of our findings. Therefore, our results should be considered exploratory and hypothesis-generating.

We enrolled 30 patients without actionable driver mutations, of whom 29 underwent surgery (96.67%). The R0 resection rate reached 93.33%, with MPR and pCR rates of 76.67% and 50.00%, respectively, surpassing our predefined endpoints. While our study demonstrated encouraging pathological response rates, suggesting potential efficacy of a shorter perioperative regimen^21, 22^, the limited small sample size and lack of a contemporaneous control group preclude definitive conclusions regarding long-term clinical benefits such as survival outcomes.

Compared with lung adenocarcinoma, sq-NSCLC is more strongly associated with smoking and predominantly affects males^23, 24^, as observed in our cohort, where only two participants were non-smoking females, while the remaining 28 were male smokers. Furthermore, common driver mutations frequently found in adenocarcinomas (such as *EGFR* alterations) are rarely identified in sq-NSCLC. Even when driver mutations are present, responses to targeted therapies and survival outcomes in sq-NSCLC remain inferior. For instance, the biomarker-driven phase II LUNG-MAP S1400 study reported an ORR of only 7% for targeted therapies in previously treated sq-NSCLC patients^23, 25–27^. High tumor mutational burden (TMB), prevalent in sq-NSCLC, is associated with a greater abundance of neoantigens and potentially enhanced antitumor immune responses^28^. In addition, sq-NSCLC tumors commonly exhibit higher PD-L1 expression and more extensive immune infiltration, both predictive of favorable immunotherapy outcomes^29^. Although several perioperative chemoimmunotherapy trials included sq-NSCLC patients, dedicated prospective trials specifically designed to evaluate perioperative immunotherapy exclusively in resectable sq-NSCLC remain relatively limited. Recent evidence indicates that sq-NSCLC may uniquely benefit from immunotherapy^30, 31^, indicating the need for subtype-specific strategies. Despite considerable advances in immunotherapy for advanced NSCLC, dedicated research on early-stage sq-NSCLC, especially in the perioperative setting, remains limited. The encouraging MPR and pCR rates observed in our study further substantiate the efficacy of immunochemotherapy in early-stage sq-NSCLC. These findings support the notion that sq-NSCLC should be recognized as a distinct subgroup within NSCLC, potentially benefiting from more tailored, histology-specific treatment strategies.

Notably, subgroup analyses for MPR and pCR did not show any significant differences based on baseline characteristics. These findings suggest that serplulimab combined with paclitaxel or nab-paclitaxel and carboplatin is broadly effective in resectable sq-NSCLC patients regardless of PD-L1 expression, consistent with the ASTRUM-004 trial^3^. Our exploratory subgroup analysis indicated no significant difference in pathological responses between patients receiving two versus three neoadjuvant cycles. However, due to limited sample size and statistical power, this observation requires validation in larger studies. Typically, standard neoadjuvant regimens for early-stage NSCLC involve three to four treatment cycles. In this study, we prospectively explored a shorter, imaging-guided neoadjuvant regimen of two to three cycles, which aligns more closely with the primary goals of neoadjuvant therapy: achieving rapid tumor shrinkage, facilitating R0 resection, and minimizing surgical morbidity. Notably, nearly all patients (96.7%) successfully underwent definitive surgery, contrasting with previous studies like the phase II NADIM trial, where approximately 10.9% of patients did not proceed to surgery after three cycles of nivolumab plus chemotherapy^32^. In other trials, about 20% of patients also failed to reach surgery^8, 9, 18–20, 33^, underscoring the advantages of a shorter regimen in reducing preoperative treatment burden. Furthermore, our innovative approach limited adjuvant treatment to one or two cycles, significantly shortening treatment duration compared to the nine to twelve months of adjuvant therapy. We hypothesize that this shorter perioperative strategy could potentially enhance patient adherence and reduce treatment burden. Nevertheless, these presumed benefits need prospective validation through patient-reported outcomes and cost-effectiveness analyses in larger randomized studies. Collectively, our findings support the clinical feasibility and efficacy of a personalized perioperative regimen with fewer cycles for resectable sq-NSCLC, encouraging further validation in phase III studies.

Interestingly, radiographic responses did not consistently align with pathological outcomes after neoadjuvant immunotherapy in our study, possibly due to mechanisms underlying immunotherapy-induced tumor responses, including enhanced immune cell infiltration and pseudo-progression. To further explore this issue, we conducted a post-hoc analysis examining changes in serum tumor markers (CEA, SCC antigen, NSE, CA125, and CYFRA21-1) during the neoadjuvant phase. Patients who had a radiographic response (CR or PR) and demonstrated a marked decrease in tumor markers were more likely to achieve a postoperative MPR. Unfortunately, due to the limited sample size, we could not draw meaningful conclusions for patients who presented with SD radiographically yet achieved MPR pathologically. Future studies with larger sample sizes are warranted to clarify the clinical utility of integrating tumor markers and radiographic assessments to optimize surgical timing and predict pathological responses. Additionally, we explored the predictive role of ctDNA clearance in identifying pCR. Our exploratory analysis indicated that ctDNA clearance correlated with pCR status, showing preliminary superiority over baseline characteristics, RECIST response, and PD-L1 expression, highlighting the potential of ctDNA monitoring as a non-invasive tool to assess the efficacy of neoadjuvant immunochemotherapy and guide subsequent therapeutic decisions. However, given the limited sample size, these findings should be validated in larger, prospective studies before clinical implementation. Our findings demonstrated that ctDNA clearance was superior to baseline characteristics, RECIST response, and PD-L1 expression in predicting pCR, highlighting the potential of ctDNA monitoring as a non-invasive tool to assess the efficacy of neoadjuvant immunochemotherapy and guide subsequent therapeutic decisions.

Consistent with our results, previous studies have shown that ctDNA levels can predict clinical outcomes in early-stage treatment-naïve NSCLC^34^ and advanced NSCLC undergoing immunotherapy^35^. Furthermore, low pretreatment levels of ctDNA have been significantly associated with improved PFS and OS in patients receiving neoadjuvant chemoimmunotherapy, and the correlation was not observed for PD-L1 expression or RECIST response ^36^. These findings suggest that ctDNA status may serve as a robust, non-invasive biomarker that accurately reflects tumor burden and treatment efficacy, potentially facilitating individualized perioperative strategies based on MRD status.

This study has several limitations. First, as a small-sample, single-arm, exploratory phase II trial without a contemporaneous control group, our results must be cautiously interpreted and viewed as hypothesis-generating. The variability in neoadjuvant treatment cycles introduces heterogeneity that limits interpretability and generalizability. Second, although not by design, all enrolled patients in this study were of Asian ethnicity. Considering the epidemiological characteristics of squamous NSCLC, our small sample size of only 30 patients inevitably resulted in a predominance of male smokers. Future studies are warranted to determine the generalizability of these results to broader patient populations. Third, while MPR is associated with improved prognosis, the absence of mature long-term survival data currently precludes definitive conclusions regarding OS benefit. Lastly, there is a potential risk of overfitting in the ctDNA analyses. Consequently, the findings regarding the association between ctDNA and pCR should be considered exploratory and hypothesis-generating. The combined prediction model is not intended for direct clinical application at this stage, but rather aims to provide insights that warrant subsequent prospective validation.

In conclusion, our study provides preliminary evidence suggesting promising efficacy and manageable safety profile of the simplified four-cycle perioperative serplulimab-based regimen in resectable sq-NSCLC. However, these findings require confirmation in larger randomized controlled trials with extended follow-up before clinical application.

## Data Availability

All data produced in the present work are contained in the manuscript.

## Resource availability

### Lead contact

Further information and requests for resources should be directed to and will be fulfilled by the lead contact, Haiquan Chen.

### Materials availability

This study did not generate new unique reagents.

### Data and code availability

All data reported in this paper will be shared by the lead contact upon request. This paper does not report original code. Any additional information required to reanalyze the data reported in this paper is available from the lead contact upon request.

## Acknowledgements

Funding: This work was supported by the National Natural Science Foundation of China (82430099 and 82573547), the Clinical Research Special Project of Shanghai Municipal Health Commission (202440070), the Medical Research Special Project for Shanghai Science and Technology Innovation Action Plan (24Y12800400), and the National Key R&D Program of China (2022YFA1103900). The study was funded by Shanghai Henlius Biotech and Burning Rock Biotech, ClinicalTrials.gov: NCT05775796.

## Author contributions

H.Q. conceptualized and designed the project. F.F., H.W., C.D., H.C., Q.H., C.Y., X.D. did the collection, analysis and interpretation of the data. F.F., H.W., C.D., H.C., Q.H., C.Y., X.D., TY, Y.Z., S.C., Y.S., J.X., S.W., YL, YZ, HH did the initial development, reviewing and editing of the manuscript. TY, Y.Z., S.C., Y.S., J.X. did the investigation and validation. H.Q. provided supervision.

All authors reviewed and approved the final version of the manuscript. All authors had full access to all the data in the study and had full responsibility for the decision to submit for publication.

## Ethics approval and consent to participate

This study was conducted in accordance with the Declaration of Helsinki and the International Conference on Harmonisation Good Clinical Practice Guidelines (ICH-GCP). The study protocol and amendments were reviewed and approved by the Clinical Research Ethics Committee of Fudan University Shanghai Cancer Center (Approval No: 2211264-18), and written informed consent was obtained from all subjects.

## Declaration of interests

The authors declare no competing interests.

## Supplemental information

Supplementary file. Figures S1-S3 and Table S1-S5

## Notes

### Competing Interest Statement

The authors have declared no competing interest.

### Clinical Trial

NCT05775796

### Author Declarations

The study protocol and amendments were reviewed and approved by the Clinical Research Ethics Committee of Fudan University Shanghai Cancer Center (Approval No: 2211264-18), and written informed consent was obtained from all participants.

