## Supplemental Tables for "Simplified Perioperative Serplulimab and Chemotherapy for Resectable Squamous NSCLC: a Phase II Trial with Biomarker Analysis"

**Supplementary Table 1.** Key Inclusion/Exclusion Criteria.

| Key Inclusion Criteria | Key Exclusion Criteria |
| --- | --- |
| Age ≥18 years and ≤ 75 years. | Other histological tumors of NSCLC include adenocarcinoma, adenosquamous carcinoma, mixed small cell carcinoma, or neuroendocrine carcinoma. |
| Histopathologically or cytologically confirmed stage II-IIIa squamous NSCLC. | Driven gene mutations, such as *EGFR* sensitivity mutations or *ALK*, *ROS1* gene rearrangements. |
| Adequate major organs function, with an expected survival of ≥12 weeks, and suitable for complete surgical resection of the pulmonary tumor as assessed by the investigator. | Confirmed CNS or other distant metastases. |
| No prior systemic antitumor therapy. | History of concurrent or active malignant disease within the past 5 years. |
| At least one measurable lesion per the RECIST version 1.1 within 4 weeks prior to the first dose. | Curative radiotherapy within 3 months prior to the first study dose. |
| ECOG PS score of 0-1. | Previous immune checkpoint inhibiting therapy. |

NSCLC, non-small cell lung cancer; RECIST, Response Evaluation Criteria in Solid Tumors; ECOG PS, Eastern Cooperative Oncology Group Performance Status; EGFR, epidermal growth factor receptor; ALK, anaplastic lymphoma kinase; ROS1, c-ros oncogene 1.

**Supplementary Table 2.** Logistic Regression Analysis of pCR and MPR in the Intention-to-Treat Population

| Variable | pCR | | MPR | |
|---|---|---|---|---|
| | OR (95% CI) | *p* | OR (95% CI) | *p* |
| Sex | | | | |
| Male | 1 | | 1 | |
| Female | 1.00 (0.04-27.00) | 1.000 | -¶ | |
| Age, years | | | | |
| <65 | 1 | | 1 | |
| ≧65 | 1.02 (0.93-1.13) | 0.622 | 0.48 (0.08-2.68) | 0.405 |
| Smoking | | | | |
| No | 1 | | 1 | |
| Yes | 1.00 (0.04-27.00) | 1.000 | -¶ | |
| ECOG PS | | | | |
| 0 | 1 | | 1 | |
| 1 | 1.00 (0.23-4.26) | 1.000 | 0.82 (0.14-4.54) | 0.818 |
| PD-L1 status* | | | | |
| Negative | 1 | | 1 | |
| Positive | 1.50 (0.21-12.99) | 0.685 | 2.53 (0.28-19.95) | 0.372 |
| Number of neoadjuvant cycles | | | | |
| 1/2 | 1 | | 1 | |
| 3 | 1.37 (0.28-6.99) | 0.691 | 1.09 (0.18-8.94) | 0.925 |
| Clinical T stage | | | | |
| T1/2 | 1 | | 1 | |
| T3/4 | 1.31 (0.31-5.63) | 0.715 | 3.25 (0.57-26.18) | 0.208 |
| Clinical N stage | | | | |
| N0 | 1 | | 1 | |
| N+ | 0.76 (0.17-3.25) | 0.713 | 0.15 (0.01-1.09) | 0.105 |
| Clinical stage | | | | |
| IIA/IIB | 1 | | 1 | |
| IIIA | 0.58 (0.13-2.46) | 0.466 | 0.82 (0.14-4.54) | 0.818 |

* PD-L1 expression status was not available for 1 pCR/MPR patient; ¶, no event, logistic regression cannot estimate parameters.

ECOG PS, Eastern Cooperative Oncology Group Performance Status; PD-L1, programmed cell death protein ligand 1.

**Supplementary Table 3.** Subgroup Analysis of pCR and MPR in the Per-Protocol Population

| Subgroup | All patients (n=29) | | | | | |
|---|---|---|---|---|---|---|
| | pCR | | | MPR | | |
| | Yes (n=15) | No (n=14) | *p* | Yes (n=23) | No (n=6) | *p* |
| Sex, n (%) | | | 1.000 | | | 1.000 |
| Male | 14 (93.33) | 13 (92.86) | | 21 (91.30) | 6 (100.00) | |
| Female | 1 (6.67) | 1 (7.14) | | 2 (8.70) | 0 (0.00) | |
| Age, years | | | 1.000 | | | 0.364 |
| <65 | 8 (53.33) | 8 (57.14) | | 14 (60.87) | 2 (33.33) | |
| ≧65 | 7 (46.67) | 6 (42.86) | | 9 (39.13) | 4 (66.67) | |
| Smoking | | | 1.000 | | | 1.000 |
| No | 1 (6.67) | 1 (7.14) | | 2 (8.70) | 0 (0.00) | |
| Yes | 14 (93.33) | 13 (92.86) | | 21 (91.30) | 6 (100.00) | |
| ECOG PS | | | 1.000 | | | 1.000 |
| 0 | 7 (46.67) | 7 (50.00) | | 11 (47.83) | 3 (50.00) | |
| 1 | 8 (53.33) | 7 (50.00) | | 12 (52.17) | 3 (50.00) | |
| PD-L1 status* | | | 1.000 | | | 0.286 |
| Negative | 2 (13.33) | 3 (21.43) | | 3 (13.04) | 2 (33.33) | |
| Positive | 12 (80.00) | 11 (78.57) | | 19 (82.61) | 4 (66.67) | |
| Number of neoadjuvant cycles | | | 1.000 | | | 0.315 |
| 1 | 0 (0.00) | 1 (7.14) | | 0 (0.00) | 1 (16.67) | |
| 2 | 10 (66.67) | 9 (64.29) | | 16 (69.57) | 3 (50.00) | |
| 3 | 5 (33.33) | 4 (28.57) | | 7 (30.43) | 2 (33.33) | |
| Clinical T stage | | | 0.715 | | | 0.169 |
| T1/2 | 7 (46.67) | 8 (57.14) | | 10 (43.48) | 5 (83.33) | |
| T3/4 | 8 (53.33) | 6 (42.86) | | 13 (56.52) | 1 (16.67) | |
| Clinical N stage | | | 1.000 | | | 0.183 |
| N0 | 7 (46.67) | 6 (42.86) | | 12 (52.17) | 1 (16.67) | |
| N+ | 8 (53.33) | 8 (57.14) | | 11 (47.83) | 5 (83.33) | |
| Clinical stage | | | 0.715 | | | 1.000 |
| IIA/IIB | 8 (53.33) | 6 (42.86) | | 11 (47.83) | 3 (50.00) | |
| IIIA | 7 (46.67) | 8 (57.14) | | 12 (52.17) | 3 (50.00) | |

* PD-L1 expression status was not available for 1 pCR/MPR patient.

ECOG PS, Eastern Cooperative Oncology Group Performance Status; PD-L1, programmed cell death protein ligand 1.

**Supplementary Table 4.** Logistic Regression Analysis of pCR and MPR in the Per-Protocol Population

| Variable | pCR | | MPR | |
|---|---|---|---|---|
| | OR (95% CI) | *p* | OR (95% CI) | *p* |
| Sex, n (%) | | | | |
| Male | 1 | | 1 | |
| Female | 0.93 (0.03-25.13) | 0.960 | -¶ | |
| Age, years | | | | |
| <65 | 1 | | 1 | |
| ≧65 | 1.02 (0.94-1.13) | 0.602 | 0.32 (0.04-2.00) | 0.240 |
| Smoking | | | | |
| No | 1 | | 1 | |
| Yes | 1.08 (0.04-29.15) | 0.960 | -¶ | |
| ECOG PS | | | | |
| 0 | 1 | | 1 | |
| 1 | 1.14 (0.26-5.02) | 0.858 | 1.09 (0.17-7.02) | 0.924 |
| PD-L1 status* | | | | |
| Negative | 1 | | 1 | |
| Positive | 1.64 (0.23-14.26) | 0.624 | 3.17 (0.34-26.49) | 0.279 |
| Number of neoadjuvant cycles | | | | |
| 1/2 | 1 | | 1 | |
| 3 | 1.25 (0.26-6.41) | 0.782 | 0.87 (0.13-7.38) | 0.891 |
| Clinical T stage | | | | |
| T1/2 | 1 | | 1 | |
| T3/4 | 1.52 (0.35-6.85) | 0.573 | 6.5 (0.87-135.35) | 0.111 |
| Clinical N stage | | | | |
| N0 | 1 | | 1 | |
| N+ | 0.86 (0.19-3.75) | 0.837 | 0.18 (0.01-1.38) | 0.148 |
| Clinical stage | | | | |
| IIA/IIB | 1 | | 1 | |
| IIIA | 0.66 (0.15-2.83) | 0.573 | 1.09 (0.17-7.02) | 0.924 |

* PD-L1 expression status was not available for 1 pCR/MPR patient; ¶, no event, logistic regression cannot estimate parameters.

ECOG PS, Eastern Cooperative Oncology Group Performance Status; PD-L1, programmed cell death protein ligand 1.

**Supplementary Table 5.** Changes in Tumor Biomarkers Following Neoadjuvant Therapy

| | Baseline (mean ± S.D.) | After Neoadjuvant (mean ± S.D.) | *p* |
|---|---|---|---|
| CEA, ng/mL | 2.55 ± 2.26 | 2.29 ± 3.02 | **0.038** |
| MPR | 2.19 ± 1.29 | 1.74 ± 1.21 | |
| CR/PR | 2.03 ± 1.06 | 1.59 ± 1.04 | |
| SD | 3.00 ± 2.13 | 2.65 ± 2.05 | |
| Non-MPR | 3.91 ± 4.30 | 4.31 ± 6.07 | |
| CR/PR | 2.26 ± 1.70 | 1.73 ± 0.11 | |
| SD | 5.56 ± 5.94 | 6.90 ± 8.49 | |
| CA125, U/mL | 24.02 ± 23.81 | 16.36 ± 12.92 | **0.034** |
| MPR | 26.76 ± 26.04 | 17.14 ± 14.17 | |
| CR/PR | 27.14 ± 27.85 | 18.32 ± 14.94 | |
| SD | 25.01 ± 18.63 | 9.66 ± 1.91 | |
| Non-MPR | 14.01 ± 7.63 | 12.95 ± 3.48 | |
| CR/PR | 18.03 ± 9.50 | 14.3 ± 3.50 | |
| SD | 9.98 ± 2.56 | 10.94 ± 3.20 | |
| CYFRA21-1, ng/mL | 6.57 ± 4.30 | 2.69 ± 1.41 | **<0.001** |
| MPR | 5.73 ± 3.94 | 2.67 ± 1.38 | |
| CR/PR | 5.77 ± 4.32 | 2.47 ± 1.22 | |
| SD | 5.53 ± 1.27 | 3.92 ± 1.99 | |
| Non-MPR | 9.82 ± 4.40 | 2.76 ± 1.67 | |
| CR/PR | 11.88 ± 4.30 | 1.72 ± 0.38 | |
| SD | 7.76 ± 4.15 | 3.80 ± 1.88 | |
| NSE, ng/mL | 15.15 ± 6.53 | 10.75 ± 2.44 | **<0.001** |
| MPR | 15.44 ± 5.63 | 11.21 ± 2.26 | |
| CR/PR | 15.91 ± 6.10 | 11.22 ± 2.45 | |
| SD | 13.25 ± 1.19 | 11.13 ± 0.12 | |
| Non-MPR | 14.01 ± 9.85 | 9.07 ± 2.49 | |
| CR/PR | 18.73 ± 13.18 | 8.87 ± 3.77 | |
| SD | 9.28 ± 1.24 | 9.26 ± 1.07 | |
| SCC Ag, ng/mL | 2.72 ± 2.80 | 1.18 ± 0.64 | **0.001** |
| MPR | 2.45 ± 2.89 | 1.26 ± 0.67 | |
| CR/PR | 2.60 ± 3.15 | 1.20 ± 0.67 | |
| SD | 1.78 ± 0.88 | 1.62 ± 0.72 | |
| Non-MPR | 3.75 ± 2.39 | 0.83 ± 0.27 | |
| CR/PR | 4.14 ± 3.04 | 0.80 ± 0.37 | |
| SD | 3.36 ± 2.14 | 0.88 ± 0.03 | |

CEA, carcinoembryonic antigen; SCC Ag, squamous cell carcinoma antigen; NSE, neuron-specific enolase; CA125, cancer antigen 125; CYFRA21-1, cytokeratin-19 fragments; MPR, major pathological response; pCR, pathological complete response; CR, complete response; PR, partial response; SD, stable disease; S.D., standard deviation.

**Supplementary Table 6. Adverse Events During the Treatment**

| AE, n (%) | Grade 1-2 | Grade 3 | Grade 4 |
|---|---|---|---|
| Any events | 30 (100.00) | 10 (26.66) | 0 |
| Aminotransferase increased | 4 (13.33) | 0 | 0 |
| Anaemia | 22 (73.33) | 0 | 0 |
| Anorexia | 6 (20.00) | 0 | 0 |
| Constipation | 6 (20.00) | 0 | 0 |
| Diarrhea | 5 (16.67) | 0 | 0 |
| Fatigue | 9 (30.00) | 0 | 0 |
| Hyperthyroidism | 3 (10.00) | 0 | 0 |
| Hypothyroidism | 4 (13.33) | 0 | 0 |
| Myalgia | 9 (30.00) | 0 | 0 |
| Myocarditis | 1 (3.33) | 1 (3.33) | 0 |
| Nausea | 4 (13.33) | 0 | 0 |
| Neutropenia | 13 (43.33) | 7 (23.33) | 0 |
| Peripheral sensory neuropathy | 3 (10.00) | 0 | 0 |
| Skin disorders | 8 (26.67) | 0 | 0 |
| Thrombocytopenia | 11 (36.67) | 0 | 0 |
