## Supplemental Figures for "Simplified Perioperative Serplulimab and Chemotherapy for Resectable Squamous NSCLC: a Phase II Trial with Biomarker Analysis"

### Supplementary Figures

**Supplementary Figure 1.** Study Design.

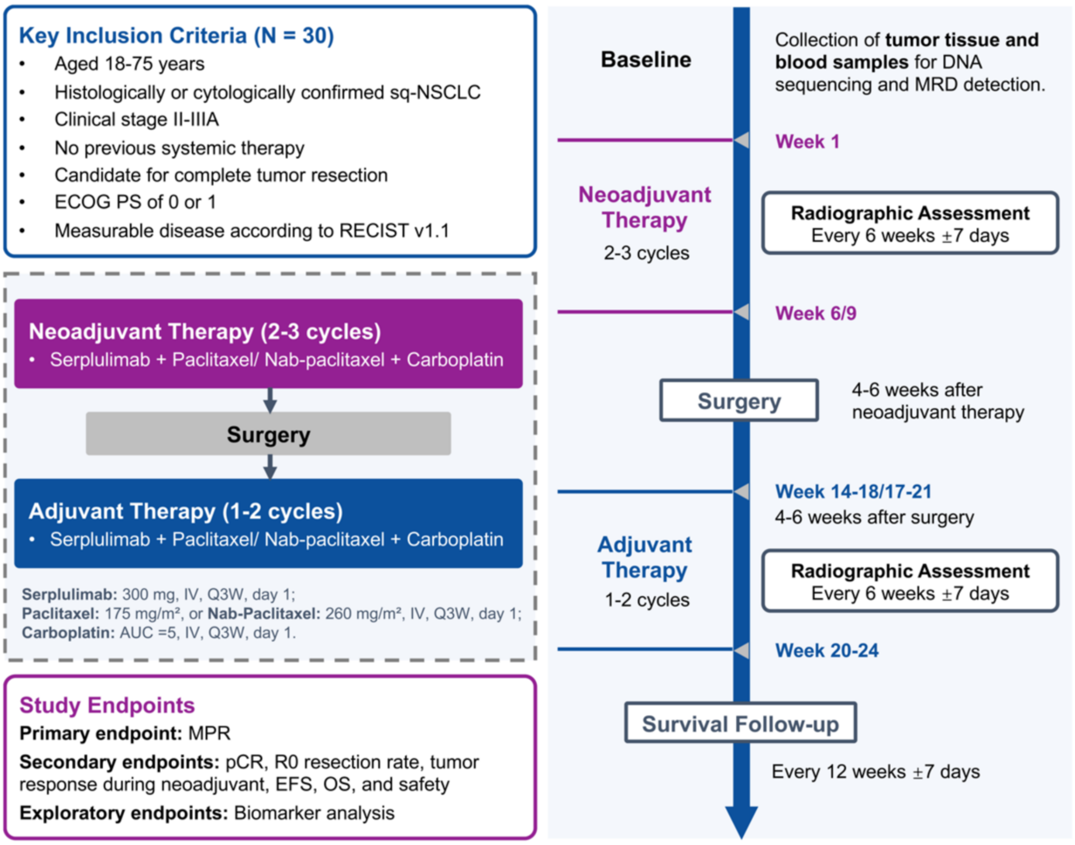


RECIST v1.1, Response Evaluation Criteria in Solid Tumors version 1.1; sq-NSCLC, squamous non-small cell lung cancer; ECOG PS, Eastern Cooperative Oncology Group Performance Status; IV, intravenous infusion; Q3W, every three weeks; AUC, area under the curve; MPR, major pathological response; pCR, pathological complete response; EFS, event-free survival; OS, overall survival; MRD, minimal residual disease.

**Supplementary Figure 2. Event-free survival and overall survival in the intention-to-treat population.**

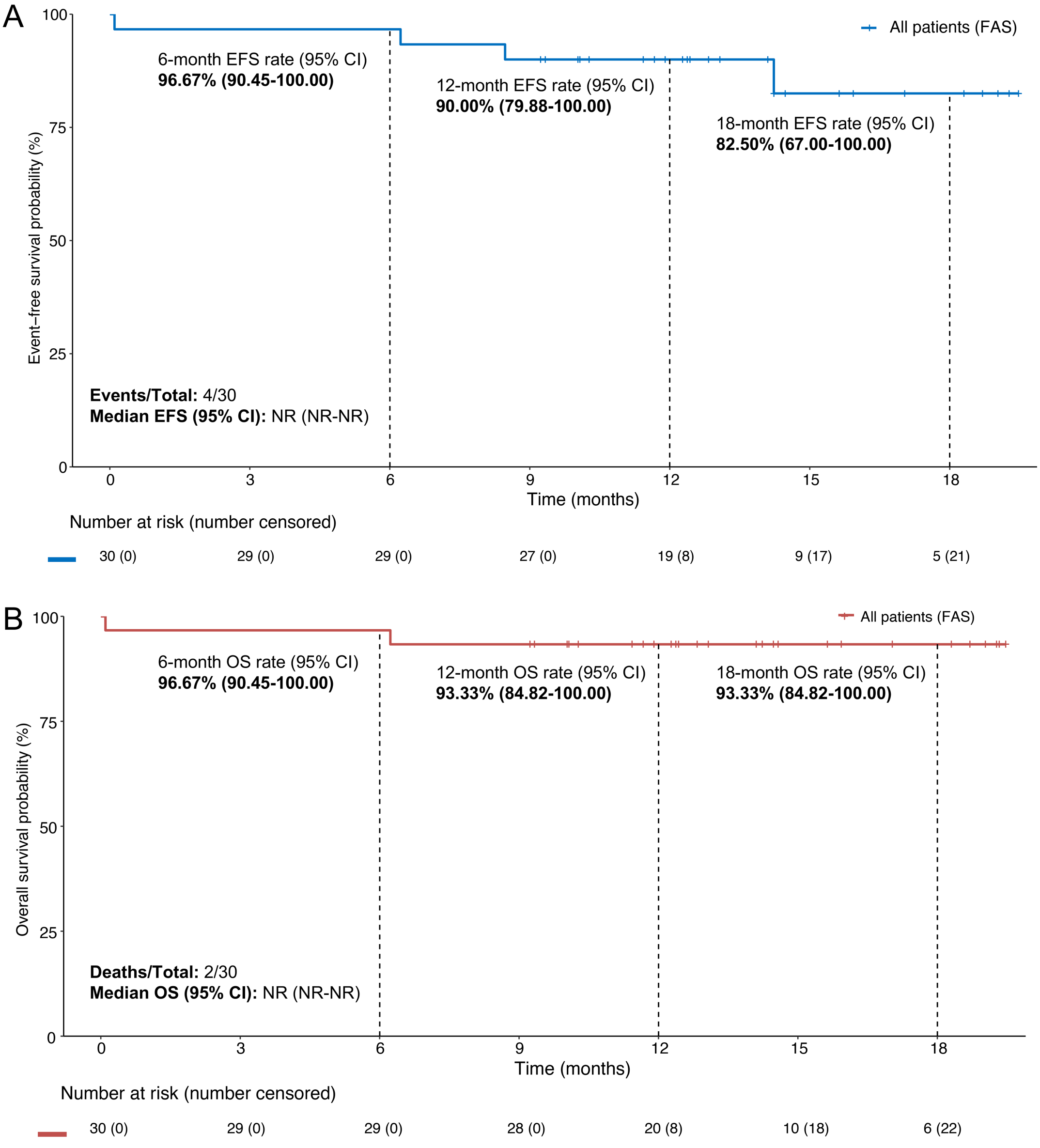


(A) Kaplan–Meier estimates of event-free survival and (B) overall survival in the overall population.

FAS, full analysis set; EFS, event-free survival; OS, overall survival.

**Supplementary Figure 3.** Event-free survival and overall survival in the per-protocol population.

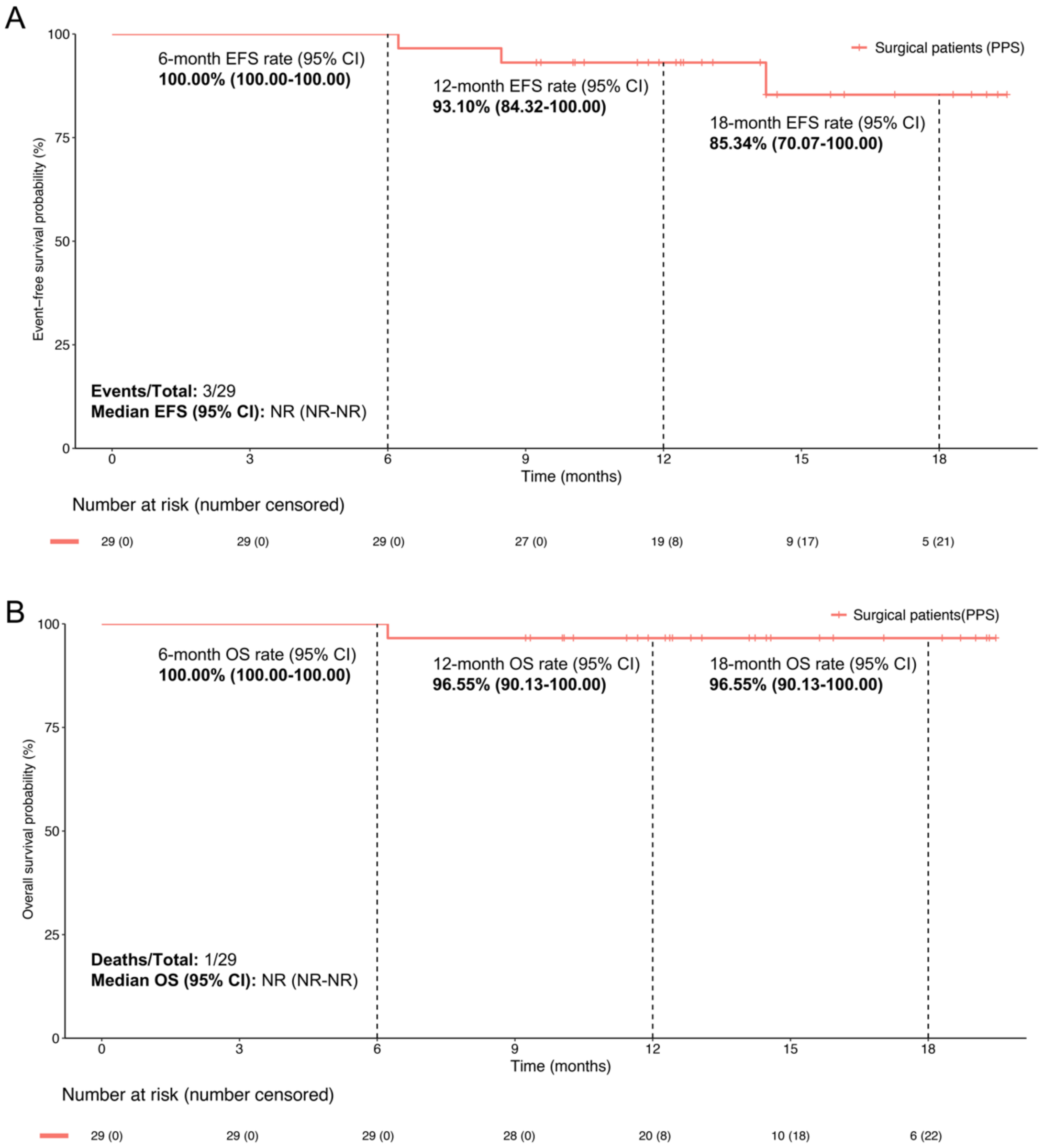


(A) Kaplan–Meier estimates of event-free survival and (B) overall survival in the per-protocol population.

FAS, full analysis set; EFS, event-free survival; OS, overall survival.

**Supplementary Figure 4.** Changes in Serum Levels of Tumor Biomarkers Following Neoadjuvant Therapy.

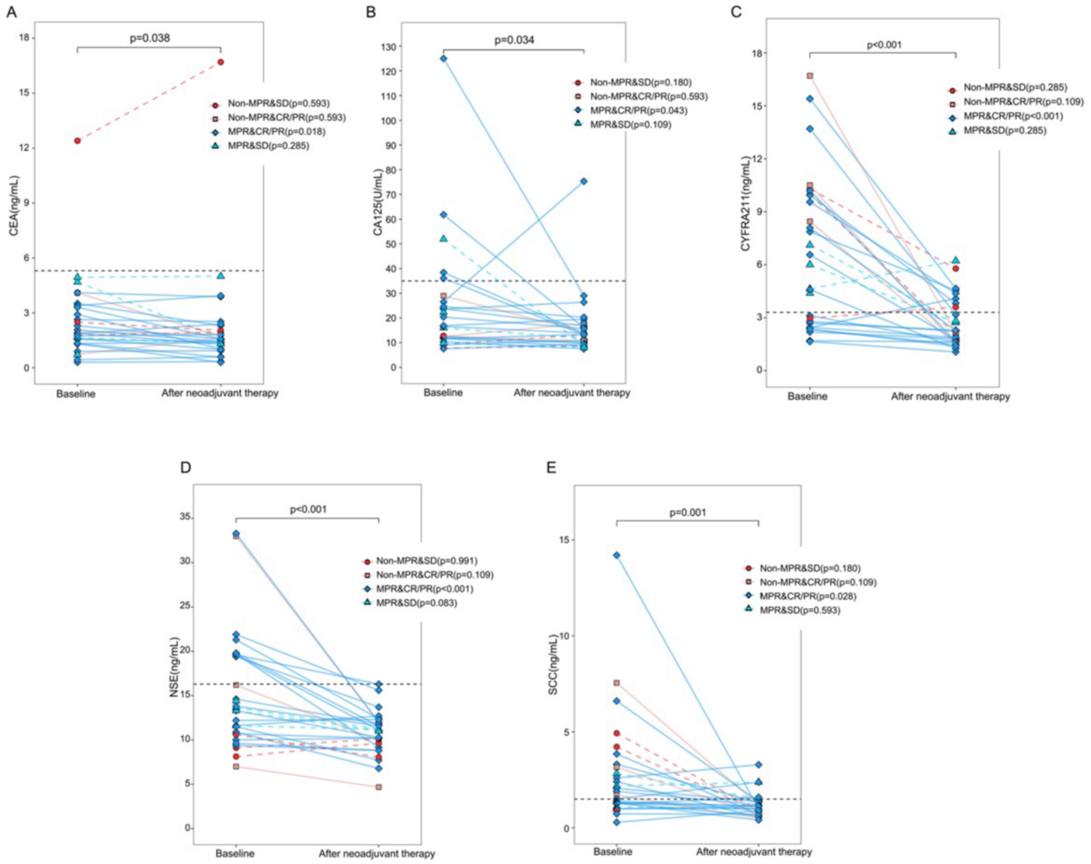


Changes in serum concentrations of the tumor markers CEA (A), CA125 (B), CYFRA21-1 (C), NSE (D), and SCC Ag (E) between baseline and post-neoadjuvant therapy (before surgery).

CEA, carcinoembryonic antigen; SCC Ag, squamous cell carcinoma antigen; NSE, neuron-specific enolase; CA125, cancer antigen 125; CYFRA21-1, cytokeratin-19 fragments; MPR, major pathological response; pCR, pathological complete response; CR, complete response; PR, partial response; SD, stable disease.
